# Chikungunya intra-vector dynamics in *Aedes albopictus* from Lyon (France) upon exposure to a human viremia-like dose range reveals vector barrier’s permissiveness and supports local epidemic potential

**DOI:** 10.1101/2022.11.06.22281997

**Authors:** Barbara Viginier, Lucie Cappuccio, Céline Garnier, Edwige Martin, Carine Maisse, Claire Valiente Moro, Guillaume Minard, Albin Fontaine, Sébastian Lequime, Maxime Ratinier, Frédérick Arnaud, Vincent Raquin

## Abstract

Arbovirus emergence and epidemic potential, as approximated by the vectorial capacity formula, depends on host and vector parameters, including the vector’s intrinsic ability to replicate then transmit the pathogen known as vector competence. Vector competence is a complex, time dependent, quantitative phenotype influenced by biotic and abiotic factors. A combination of experimental and modelling approaches is required to assess arbovirus intra-vector dynamics and estimate epidemic potential. In this study, we measured infection, dissemination, and transmission dynamics of chikungunya virus (CHIKV) in a field-derived Aedes albopictus population (Lyon, France) after oral exposure to a range of virus doses spanning human viraemia. Statistical modelling indicates rapid and efficient CHIKV progression in the vector mainly due to an absence of a dissemination barrier, with 100% of the infected mosquitoes ultimately exhibiting a disseminated infection, regardless of the virus dose. Transmission rate data revealed a time-dependent, but overall weak, transmission barrier, with individuals transmitting as soon as 2 days post-exposure (dpe) and >50% infectious mosquitoes at 6 dpe for the highest dose. Based on these experimental intra-vector dynamics data, epidemiological simulations conducted with an agent-based model showed that even at low mosquito biting rates, CHIKV could trigger outbreaks locally. Together, this reveals the epidemic potential of CHIKV upon transmission by Aedes albopictus in mainland France.

## Introduction

Arthropod-borne viruses (arboviruses) are pathogens transmitted to vertebrate hosts by hematophagous arthropods, mainly mosquitoes. Arbovirus spread is a multi-factorial, dynamic process that can be estimated using the vectorial capacity (VCap) model, which aims to determine the average number of infectious vector bites that arise per day from one infected host in a susceptible human population (Smith et al., 2012). The vector-centric component of VCap integrates mosquito ecological (density per host, survival) and behavioural (daily biting rate per human) factors along with the vector’s proxies of virus transmission efficiency such as vector competence (VComp) and its time-related expression, the extrinsic incubation period (EIP). VComp represents the ability of mosquitoes to : i) allow midgut infection following an infectious blood meal, ii) disseminate the virus beyond the midgut barrier, and iii) retransmit the virus through the saliva during the next bite. In VCap models, VComp and EIP are often simplified under the EIP50, the time required to reach 50% of infectious mosquitoes. Effectively, each individual mosquito has a given EIP leading to a range of EIPs in the population. EIP distribution can be assessed experimentally by measuring the time between the initial mosquito infection and the mosquito infectiousness using an adequate experimental design (transmission assay for a relevant number of individual mosquitoes and time points). Taking into account the time-dependency of Vcomp improves VCap estimation and therefore allows to capture the full epidemic potential of arboviruses (Lequime et al., 2020). VComp is impacted by biotic (*e.g.*, mosquito and virus genotype, virus dose, mosquito microbiota) and abiotic (*e.g.*, temperature) factors (Viglietta et al., 2021), but how these factors shape VComp dynamics has still to be determined.

Dengue virus (DENV), yellow fever virus (YFV), Zika virus (ZIKV), and chikungunya virus (CHIKV) pose a major sanitary threat as they are responsible for hundreds of millions of human infections each year worldwide, leading to severe morbidity and mortality (Labeaud et al., 2011; Bhatt et al., 2013). These arboviruses are primarily transmitted to humans by *Aedes aegypti* mosquitoes, although the Asian tiger mosquito, *Aedes albopictus,* is often incriminated as a vector. Indeed, *Ae. albopictus* is an important vector of arboviruses as evidenced by vector competence laboratory assays and the detection of infected field specimens (Gratz, 2004; Paupy et al., 2009). Notably, *Ae. albopictus* was identified as the main vector during CHIKV outbreaks in Gabon (2007), Congo (2011) as well as in a major outbreak at La Réunion island (2006) (Schuffenecker et al., 2006; Bonilauri et al., 2008; Pagès et al., 2009; Paupy et al., 2012; Mombouli et al., 2013). In Europe, this vector species is incriminated for autochthonous circulation of CHIKV for instance in Italy (Venturi et al., 2017) and mainland France (Delisle et al., 2015). Vector competence studies established that vector competence of *Ae. albopictus* for CHIKV depends on genetic (*e.g.*, mosquito and virus genotype) and environmental (*e.g.*, temperature) factors (Tsetsarkin et al., 2007; Vazeille et al., 2007; Zouache et al., 2014; Sanchez-Vargas et al., 2019). Host viremia, approxymated by virus dose in the blood meal during artifical mosquito infectious feeding experiments, is another major factor that drives mosquito vector competence (Nguyet et al., 2013; Aubry et al., 2020). Vertebrate host viremia for CHIKV last 4 to 12 days with an increase in blood viral titer prior to symptoms appearance, up to a peak around 8 log10 infectious particles/mL followed by a decrease until virus clearance for most of the cases (Schwartz & Albert, 2010). Beyond non-human primates, an estimate of CHIKV human viremia dynamics is lacking due to limited longitudinal monitoring of infected patients, despite it could help to decipher the duration and magnitude of human infectiousness for mosquitoes (Labadie et al., 2010). In *Ae. albopictus*, two studies exposed mosquitoes to a range of CHIKV doses in the blood meal with varying outcomes on vector competence as estimated by mosquito infection and dissemination rate (Pesko et al., 2009; Hurk et al., 2010). Vector competence studies on *Ae. albopictus* from mainland France measured CHIKV transmission rate although upon exposure to a single dose, always above 6.5 log10 FFU/mL range which does not cover the range of human vireamia (Moutailler et al., 2009; Vega-Rua et al., 2013; Zouache et al., 2014; Vega-Rúa et al., 2015). In addition, these studies focused on a limited number of time points after CHIKV exposure that prevent to capture the dynamics of vector competence. A study monitoring intra-vector dynamics of CHIKV and its epidemiological relevance is still lacking, notably upon variations of vector competence major drivers such as virus dose.

Here, we studied intra-vector infection dynamics of a field-derived population of *Ae. albopictus* from Lyon metropolis exposed by artificial membrane feeding to a range of human viraemia-like CHIKV (La Réunion 06.21 isolate, East Central South Africa (ECSA) clade) doses, based on our model of human CHIKV viremia in the blood. Strains from ECSA clade carrying the same A226V mutation on enveloppe *E1* gene than 06.21 were identified from autochtonous cases in mainland France, supporting the choice of the 06.21 strain for this study (Franke et al., 2019). Individual mosquitoes were analyzed from day 2 to day 20 post-exposure (dpe) to determine infection, dissemination, and transmission rates by infectious titration in addition to the quantification of CHIKV RNA load in the saliva. This allowed us to estimate CHIKV intra-vector dynamics and the strength of vector infection, dissemination, and transmission barriers as well as the distribution of EIP according to the virus dose in the blood meal. These data were implemented in the agent-based model *nosoi* (Lequime et al., 2020) to estimate, using realistic vectorial capacity parameters, the epidemic potential of CHIKV in a French population of *Ae. albopictus*. Our results improve our understanding on vector-virus interactions and provides key informations to better anticipate and prevent CHIKV emergence in mainland France.

## Methods

### Modelling chikungunya viraemia in humans

Chikungunya virus (CHIKV) loads in human blood along with the time course of infection in patients were recovered from two studies. The first study monitored blood CHIKV viraemia from a retrospective cohort of 102 febrile patients in Bandung, West Java, Indonesia, between 2005 and 2009 (Riswari et al., 2015). The second study assessed CHIKV RNA viremic profile from 36 sera from day 1 to day 7 of illness during a CHIKV epidemic in 2009 in Thepa and Chana districts of Songkhla province, Thailand (Appassakij et al., 2013). For the second study, the median value of the blood CHIKV RNA loads from a group of patients (n=2 to 21) per day of illness was used. The viraemia data from RT-qPCR was expressed on the logarithmic scale to the base 10 before model fitting. Wood’s gamma-type function was used to model the viraemia dynamic. The function is given in the following equation:

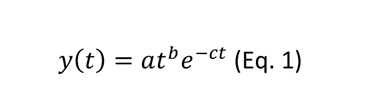

where *y(t)* represents the level of viraemia in the blood at *t* days post-infection, with *a*, *b,* and *c* representing constants linked to the viraemia dynamics (Islam et al., 2013). Viraemia data were originally expressed in time pre-or post-symptom onset, while the model represents viraemia as a function of time post-infection. A fixed arbitrary median intrinsic incubation period of 6 days was added to each viraemia time to standardize the time scale between the data and the model. This fixed incubation period falls into the estimated 2-10 days incubation range (Moloney et al., 2014) and was chosen to ensure that all observed viraemia data occurred after infection. The model was fitted to the data using non-linear least-squares regression implemented in the *nls* function in the R environment (RCoreTeam, 2022). This method proposed a possible intra-human CHIKV viraemia dynamic with 95% confidence intervals.

### CHIKV stock production and titration

The CHIKV strain 06.21 from the Indian Ocean lineage was isolated from a newborn serum sample with neonatal encephalopathy in La Réunion island in 2005 (Schuffenecker et al., 2006). This strain was amplified in *Aedes albopictus* cell line C6/36 as previously described (Raquin et al., 2015). CHIKV was inoculated at a multiplicity of infection of 0.01 on *Ae. albopictus* C6/36 cells cultivated in Leibovitz’s L-15 media (Gibco) with 10 % (v:v) 1X Tryptose Phosphate Broth (Gibco), 10 % (v:v) foetal bovine serum and 0.1 % (v:v) 10,000 units/mL penicillin/streptomycin (Gibco). Cells were incubated for 3 days at 28 °C before the cell supernatant was clarified by centrifugation for 5 min at 500 g and stored at −80 °C as aliquots. CHIKV infectious titer was measured on C6/36 using fluorescent focus assay (Raquin et al., 2015). Briefly, 3×10^5^ cells/well were inoculated in 96-well plates (TPP) with 40 µL/well of viral inoculum (after culture media removal) and incubated for 1 h at 28 °C. 150 µL/well of a mix 1:1 L-15 media and 3.2% medium viscosity carboxymethyl cellulose (Sigma) were added as an overlay before incubation of the cells for 3 days at 28 °C. After incubation, cells were fixed in 150 µL/well of 4 % paraformaldehyde for 20 min at room temperature (RT) and then rinsed 3 times in 100 µL/well of 1X Dulbecco’s phosphate-buffered saline (DPBS) (Gibco) prior to immune labelling. Cells were permeabilized for 30 min in 50 µL/well of 0.3 % (v:v) Triton X-100 (Sigma) in 1X DPBS + 1 % Bovine Serum Albumin (BSA, Sigma) at RT then rinsed 3 times in 100 µL/well of 1X DPBS. A Semliki Forest virus anticapsid antibody cross-reacting with CHIKV was used as a primary antibody, diluted 1:600 in 1X DPBS + 1 % BSA (Greiser-Wilke et al., 1989). Cells were incubated in 40 µL/well of primary antibody for 1 h at 37°C, rinsed 3 times in 100 µL/well of 1X DPBS then incubated in 40 µL/well of anti-mouse Alexa488 secondary antibody (Life Technologies) at 1:500 in 1X DPBS + 1 % BSA for 30 min at 37 °C. Cells were rinsed 3 times in 100 µL/well 1X DPBS, then once in 100 µL/well tap water, stored at 4 °C overnight before the enumeration of fluorescent foci under Zeiss Colibri 7 fluorescence microscope at 10X objective. The CHIKV infectious titer was expressed as the log10 fluorescent focus unit (FFU) per mL. Plates were then stored at 4 °C protected from light to allow further reading. The infectious titer of the neat CHIVK 06.21 stock was 8.63 log10 FFU/mL.

### Mosquito colony maintenance

The Lyon metropolis population of *Aedes albopictus* originates from a field sampling of larvae in 2018 that were brought back to insectary for rearing (Microbial Ecology lab, Lyon, France). Sampling locations included Villeurbanne (N : 45°46’18990’’ E : 4°53’24615’’) and Pierre-Bénite (N :45°42’11534’’ E : 4°49’28743’’) in Lyon metropolis area, mainland France. Mass rearing of the population under standard laboratory conditions (28 °C, 80% relative humidity, 16:8 hours light:dark cycles) using mice feeding (*Mus musculus*) allowed to maintain genetic diversity, in accordance with the Institutional Animal Care and Use Committee from Lyon1 University and the French Ministry for Higher Education and Research (Apafis #31807-2021052715018315). Prior to infectious blood feeding, eggs were hatched for 1 h in dechlorinated tap water, and larvae were reared at 26 °C (12:12h light:dark cycle) at a density of 200 larvae in 23 x 34 x 7 cm plastic trays (Gilac) in 1.5 L of dechlorinated tap water supplemented with 0.1 g of a 3:1 (TetraMin tropical fish food:Biover yeast) powder every two days. Adults were maintained in 32.5 x 32.5 x 32.5 cm mesh cages (Bugdorm) at 28 °C, 80 % relative humidity, 12:12h light:dark cycle with permanent access to 10% sugar solution.

### Experimental mosquito exposure to CHIKV

Female mosquitoes (4 to 8-day old) from F10 generation were confined in 136 x 81 mm plastic feeding boxes (Corning-Gosselin) with ∼60 individual per box then transferred to the level 3 biosafety facility (SFR Biosciences, AniRA-L3, Lyon Gerland) at 26 °C, 12:12 h light:dark cycle deprived from sugar solution 16 h before the infectious blood meal. The blood meal was composed of a 2:1 (v:v) mixture of washed human erythrocytes (from multiple anonymous donors collected by EFS AURA under the CODECOH agreement DC-2019-3507) and viral suspension at several doses, and supplemented with 2 % (v:v) of 0.5 M ATP, pH 7 in water (Sigma). Feeders (Hemotek) were covered with pig small intestine and filled with 3 mL of infectious blood mixture. Females were allowed to feed for 1 h at 26 °C and blood aliquots were taken before (T0) and after (1h) the feeding and stored at −80 °C for virus titration (Figure S1). Mosquitoes were anaesthetized on ice and fully engorged females were transferred in 1-pint cardboard containers (10-25 females/container) and maintained with 10 % sucrose. Cardboard containers were placed in 18 x 18 x 18 inches cages (BioQuip) and kept in climatic chambers at 26 °C, 70 % humidity. Two independent vector competence experiments were conducted with 370 and 418 individuals mosquitoes per experiment, respectively. In a first experiment (n=370), mosquito body and head infection were tested for the presence of infectious virus at 4 time points while in the second experiment (n=418), mosquito head and saliva were analyzed at 10 time points.

### Mosquito dissection and CHIKV detection

At the selected day post-exposure (dpe) to CHIKV, individual saliva were collected then the heads and bodies were recovered. Prior to saliva collection, mosquitoes were anaesthetized on ice then legs and wings were removed under a stereomicroscope. Individuals were placed on plastic plates maintained by double-sided adhesive tape. The proboscis was inserted in a trimmed 10 µL filtered tip containing 10 µL of foetal bovine serum (FBS) held above the mosquito by modelling clay (Heitmann et al., 2018). Two µL of 1 % pilocarpine hydrochloride (Sigma) supplemented with 0.1 % Tween-20 (Sigma) in water were added on the thorax of each mosquito to enhance salivation. Mosquitoes were allowed to salivate at 26 °C, 80 % relative humidity for 1 h. The FBS that contains the saliva was expelled in an ice-cold tube filled with 150 µL of DMEM media (Gibco) supplemented with antibiotics solution (Amphotericin B 2.5 µg/ml, Nystatin 1/100, Gentamicin 50 µg/ml, Penicillin 5 U/ml and Streptomycin 5 µg/ml (Gibco)). Following salivation, each mosquito’s head and body were separated using a pin holder with 0.15 mm minutien pins (FST). Heads and bodies were transferred in individual grinding tubes (Qiagen) containing 500 µL of DMEM supplemented with antibiotics (see above) and one 3-mm diameter tungsten bead (Qiagen). Samples were ground on a 96-well adapter set for 2 x 1 min, 30 Hz using a TissueLyser II (Qiagen), then stored at −80 °C. CHIKV detection was performed once on 40 µL of undiluted (raw) saliva, head and body samples using fluorescent focus assay on C6/36 cells (see above). Each mosquito sample was declared positive or negative for CHIKV in the presence or absence of a fluorescent signal, respectively. Each 96-well plate contained positive (virus stock) and negative (raw grinding media) controls. Two independent persons examined each plate. Of note, saliva samples were deposited immediately (no freezing step) on C6/36 cells to maximize CHIKV detection. 30 µL of saliva sample were immediately mixed with 70 µL of TRIzol (Life Technologies) and stored at −80°C before RNA isolation. The rest of the samples were stored at −80 °C as a backup.

### RNA isolation from saliva

Total RNA was isolated from 30 µL of saliva mixed with 70 µL TRIzol and then stored at −80°C, as described (Raquin et al., 2017). After thawing samples on ice, 20 µL chloroform (Sigma) were added. The tubes were mixed vigorously, incubated at 4 °C for 5 min and centrifuged at 17,000 G for 15 min, 4 °C. The upper phase was transferred in a new tube containing 60 µL isopropanol supplemented with 1 µL GlycoBlue (Life Technologies). Samples were mixed vigorously and stored at −80 °C overnight to allow RNA precipitation. After 15 min at 17,000 G, 4 °C, the supernatant was discarded, and the blue pellet was rinsed with 500 µL ice-cold 70 % ethanol in water. The samples were centrifuged at 17,000 G for 15 min, 4 °C, the supernatant was discarded, and the RNA pellet was allowed to dry for 10 min at room temperature. Ten µL RNAse-free water (Gibco) were added, and samples were incubated at 37 °C for 10 min to solubilize RNA prior to transfer in RNAse-free 96-well plates and storage at −80 °C.

### CHIKV RNA load quantification in saliva

Total RNA (2 µL) isolated from individual mosquito saliva were used as template in a one-step TaqMan RT-qPCR assay. The QuantiTect Virus kit (Qiagen) was used to prepare the reaction mix in a final volume of 30 µL. The reaction solution consisted of 6 µL 5X master mix, 1.5 µL primers (forward 5’-CCCGGTAAGAGCGGTGAA-3’ and reverse 5’-CTTCCGGTATGTCGATGGAGAT-3’) and TaqMan probe (5’-6FAM-TGCGCCGTAGGGAACATGCC-BHQ1-3’) (Hurk et al., 2010) mixed at 0.4 µM and 0.2 µM final concentration respectively, 0.3 µL 100X RT mix, 20.2 µL RNAse-free water (Gibco) and 2 µL template saliva RNA. RT-qPCR reaction was conducted on a Step One Plus machine (Applied) for 20 min at 50 °C (RT step), 5 min at 95°C (initial denaturation) and 40 cycles with 15 s at 95 °C and 45 s at 60 °C. Serial dilutions of CHIKV 06.21 synthetic RNA from 8 to 1 log10 copies/µL were used as an external standard to estimate CHIKV RNA copies in saliva samples. Each plate contained duplicates of standard synthetic RNA samples as well as negative controls and random saliva samples without reverse transcriptase (RT-) also in duplicate. Aliquots from the same standard RNA (thawed only once) were used for all the plates, and samples from a single time-point were measured on the same plate to allow sample comparison.

### Statistical analyses

Mosquito infection (number of CHIKV-positive mosquito bodies / number engorged mosquitoes), dissemination (number of CHIKV-positive heads / number of CHIKV-positive bodies) and transmission rate (number of positive CHIKV-saliva / number of CHIKV-positive heads) were analysed by logistical regression and considered as binary response variables. The time (dpe) and virus dose (log10 FFU/mL) were considered continuous explanatory variables in a full factorial generalized linear model with a binomial error and a logit link function. Logistic regression assumes a saturation level of 100% and could not be used to model the relationship between the probability of transmission (response variable) and the time post-infection, the dose and their interaction (predictors). Therefore, we first estimated the saturation level (K) for each dose and subtracted the value N = number of mosquitoes with CHIKV dissemination x (100% - K) to the number of mosquitoes without virus in their saliva at each time post virus exposure to artificially remove mosquitoes that would never ultimately transmit the virus from the dataset. Logistic regression was then used on these transformed data to predict transmission rates across time post virus exposure and the virus dose (Figure S2). The statistical significance of the predictors’ effects was assessed by comparing nested models using deviance analysis based on a chi-squared distribution. All the statistical analyses were performed in R Studio (Posit), and figures were created with the package *ggplot2* within the *Tidyverse* environment (Wickham et al., 2019). The R script used for this study is available, and supplementary table 1 summarizes the proportion of infected, disseminated and infectious mosquitoes for all the experiments conducted.

### Epidemiological modelling using *nosoi*

A series of stochastic agent-based model simulations were performed using the R package *nosoi* and a specific branch available on *nosoi*’s GitHub page (https://github.com/slequime/nosoi/tree/fontaine) as previously done (Lequime et al., 2020). Briefly, 100 independent simulations were run in replicates for each condition. Each simulation started with one infected human and was run for 365 days or until the allowed number of infected individuals (100,000 humans or 1,000,000 mosquitoes, respectively) was reached. We considered transmission only between an infected mosquito and an uninfected human or between an infected human and an uninfected mosquito. Vertical and sexual transmission, and the impact of potential superinfection were ignored during the simulations. We assumed no particular structure within host and vector populations.

It was also considered that humans do not die from infection and leave the simulation after they clear the infection (here 12 days). Each human agent experienced a Poisson-like distribution of bites per day with a mean value manually set at 1, 2, 3, 4, 5, 6, 7, 8, 9, 10 or 60 based on field measurement of *Aedes albopictus* blood-feeding behaviour (Delatte et al., 2010). Human-to-mosquito transmission followed a time-post-infection-dependent probability function, identical for all human agents, computed from the human viraemic profile (Eq. 1, see above) and dose-response experiments (Eq. 2) :

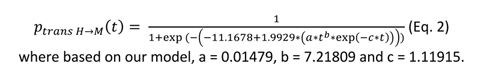

The daily survival probability of infected mosquito agents was set empirically at 0.85 (Favier et al., 2005; Fontaine et al., 2018). Human biting (only one per mosquito agent) was set at fixed dates depending on a gonotrophic cycle duration drawn for each mosquito in a truncated Poisson distribution with a mean of 4 days (no draws below 3). The mosquito-to-human transmission was determined for each mosquito agent based on its individual EIP value acting as a threshold for transmission (if time post-infection is greater or equal to the EIP value, the mosquito can transmit). However, a certain proportion (based on saturation parameter K, see above) of mosquitoes never transmitted. The individual EIP value was dependent on the virus dose that initiated the infection based on this equation (Lequime et al., 2020):

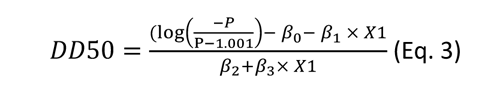

where P = 0.5 (i.e., the median transmission probability), *β0* is the Y-intercept value (−2.328973), *β1* (0.278953), *β2* (0.136746) and *β3* (0.003276) are model coefficients associated to the virus dose, time post virus exposure and their interaction, respectively. X1 represents the virus dose value.

## Results

### Estimating CHIKV viraemia in humans by modelisation of clinical data

Intra-human dynamic of CHIKV viraemia over time post-infection was approximated using time course of human CHIKV viraemia in individual patients from two studies (Appassakij et al., 2013; Riswari et al., 2015). CHIKV loads were assessed at 3 to 6 different time points, prior or post symptoms onset from the blood of 5 patients. The range of CHIKV viraemia duration among the 5 patients was 4-12 days, with a minimal and a maximum CHIKV load of 1 and 8.78 log10 PFU equivalent/mL, respectively. Of note, two patients displayed 1.04 and 3.25 log10 PFU equivalent/mL before symptoms onset, respectively. Modelling CHIKV viraemic profile using a Wood’s gamma-type function indicates that mean viral load rapidly increases to peak after 6.45 days (*i.e.* within 24 h after symptoms onset) at 7.55 log10 PFU equivalent/mL (7.01-8.78 log10 equivalent PFU/mL depending on the patient) (Figure 1).

**Figure 1.**
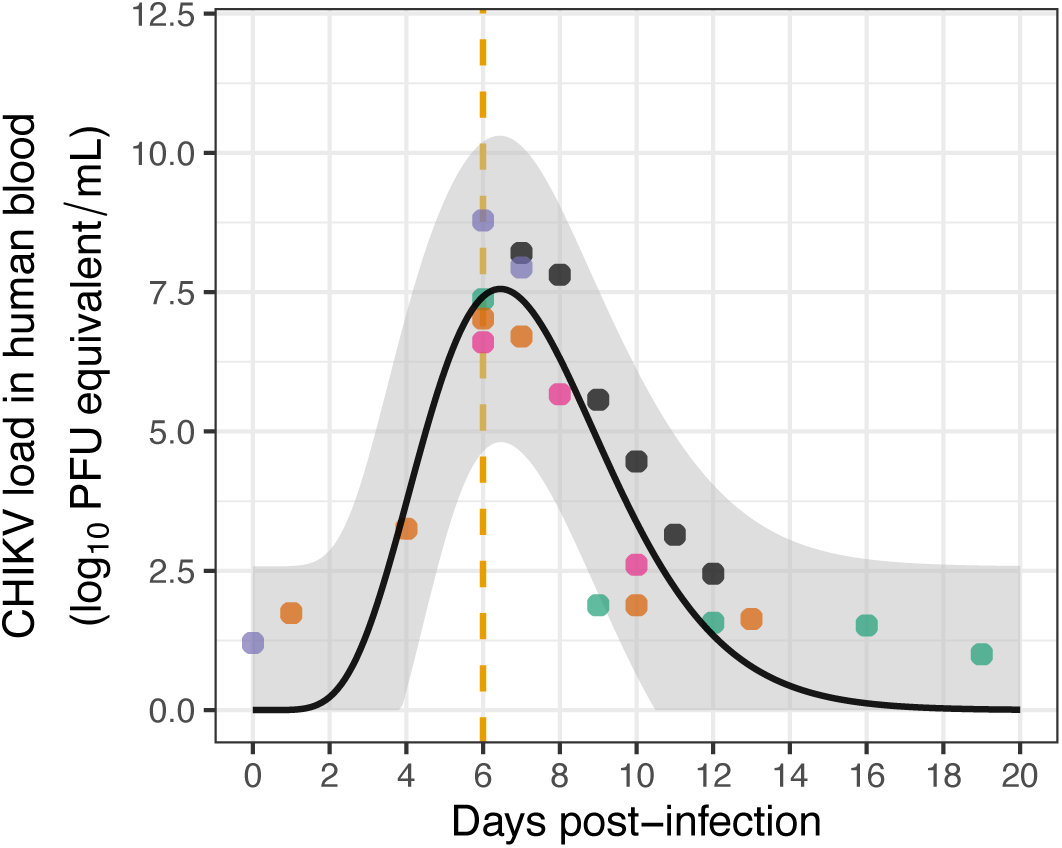
Estimated time course of CHIKV load in human blood as a function of days post-infection. A Wood’s gamma-type function was used to model CHIKV viraemia dynamics based on the time course of human viraemia data in 5 patients. The black line represents model prediction using mean fit parameter values. Each dot represents a single experimental measurement with colours corresponding to different patients. The vertical gold line indicates the day of symptoms onset. The grey ribbon represents upper and lower predicted values. Refers to raw data table “ChikV_Viremia_dynamic.txt” (see data availability section).

### The dose, but not the time, modulates mosquito infection rate

Female *Ae. albopictus* were seprately exposed to a human erythrocytes suspension containing three CHIKV doses (3.94, 6.07 and 8.63 log10 FFU/mL) that span the estimated range of human viraemia as estimated above (Figure 1). The mortality rate was very low regardless of time or oral dose, remaining below 5%. Mosquito infection rate (IR) remained below 7 % (n=35 to 53 individuals tested) at 3.94 log10 FFU/mL, ranged from 65 to 85 % at 6.07 log10 FFU/mL (n=20 to 35) and raised above 94 % at 8.63 log10 FFU/mL (n=12 to 24) (Figure 2A). A detail table of these data is presented in supplementary table S1. IR significantly increases with the dose but it does not depend on the time post-exposure (Wald χ^2^, *P*dose = 1.1 x 10^-6^, *P*time = 0.9 and *P*dose*time = 0.17). As IR depends on the virus dose but not on the time post-exposure, we fitted a logistic model to the data considering CHIKV titer in the blood meal as a unique explanatory variable (Figure 2B). Dose-dependent IR describes a sigmoid with a median oral infection dose (OID50%) of 5.6 log10 FFU/mL and an OID25% and OID75% of 5.05 log10 FFU/mL and 6.15 log10 FFU/mL, respectively. The oral infection saturation level was reached at about 7.5 log10 FFU/mL.

**Figure 2.**
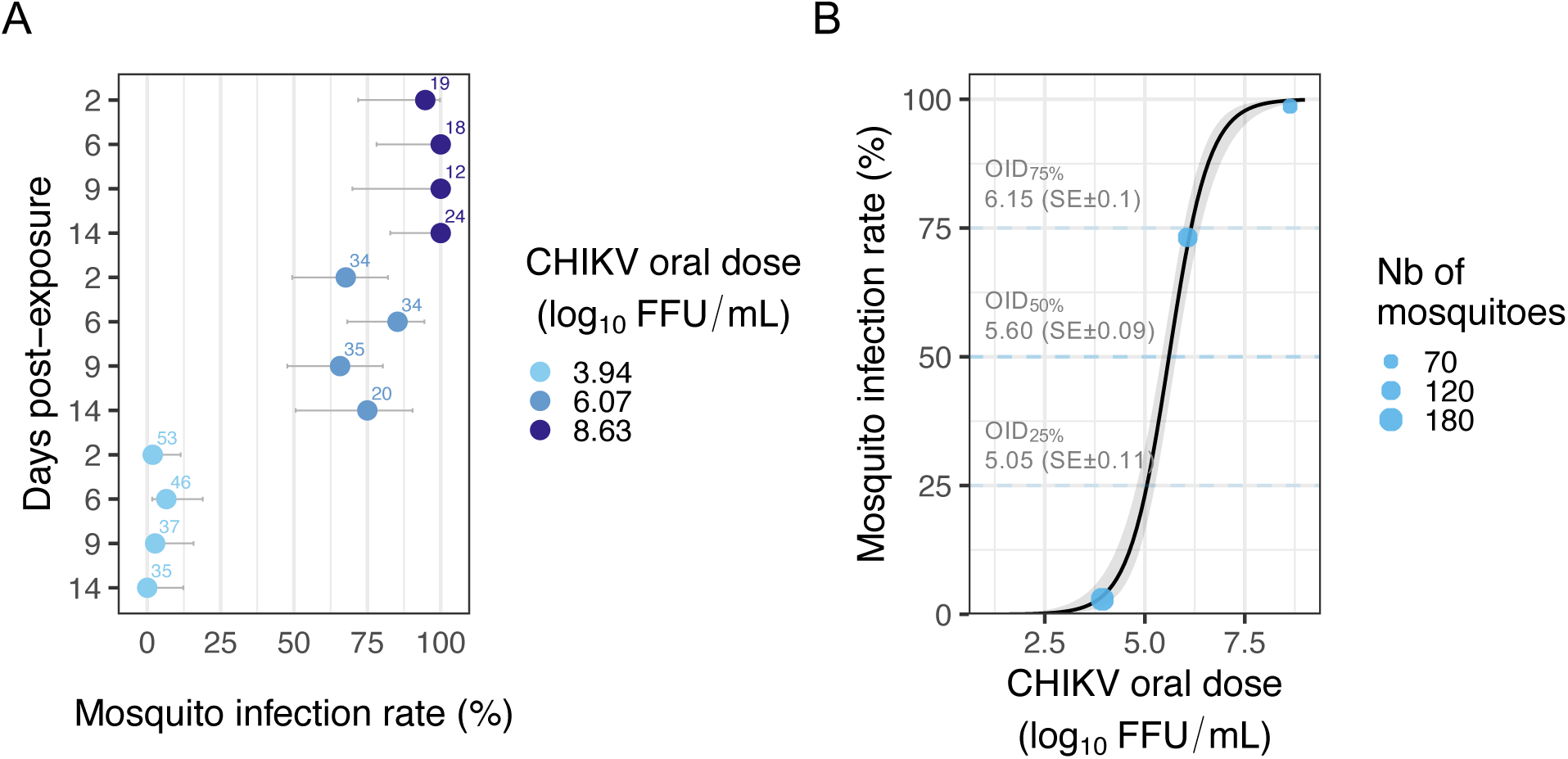
Dose-dependent infection rate of *Ae. albopictus* mosquitoes exposed to CHIKV 06.21. (A) The mosquito infection rate corresponds to the proportion (in %) of mosquito bodies positive for CHIKV infection out of the total of engorged mosquitoes, measured at 2, 6, 9 and 14 days post-exposure for three CHIKV doses (3.94, 6.07 and 8.63 log10 FFU/mL) in the blood meal. The number of individuals analysed at each time point is indicated above the bars that represents the 95% confidence interval. (B) Mosquito infection rate as a function of CHIKV dose in the blood meal. Blue dots correspond to the observed infection rate upon the three CHIKV doses tested. Dot size is proportional to the number of mosquitoes tested. The black line was obtained by fitting a logistic model to the data. The grey ribbon indicates the 95 % confidence interval. The oral infectious dose (OID) to infect 25 %, 50 % and 75 % of the mosquitoes exposed to CHIKV is indicated with the associated standard error (in log10 FFU/mL). Refers to raw data table “Data_titer_EIPdyna_body_head_final.txt” (see data availability section).

### Dose-and time-dependent mosquito dissemination dynamics

The proportion of CHIKV-positive heads among positive bodies (*i.e.,* mosquito dissemination rate, DIR) was analysed using virus dose, time post-exposure and their interaction as explanatory variables. At 2 days post-exposure, <50 % of the mosquitoes presented a disseminated infection for the doses 3.94 (n=1 individual tested) and 6.07 (n=18) log10 FFU/mL, whereas DIR was already above 80 % after 2 days in mosquitoes exposed to 8.63 log10 FFU/mL of CHIKV (n=23) (Figure 3A). Notably, DIR increases >80 % for all the three doses after 6 dpe (n=1 to 29 individual tested per time point and dose) (Figure 3A and Table S1). Although they are not in interaction, both time and dose impact DIR (Wald χ^2^, *P*dose = 8.29 x 10^-6^, *P*time = 1.75 x 10^-6^ and *P*dose*time = 0.83). The plateau was 100 % at doses 3.94 and 8.63 log10 FFU/mL and 95.6% at the dose 6.07 log10 FFU/mL (n=22/23). The time to reach 50 % dissemination in *Ae. albopictus* exposed to CHIKV was 7.5 days, 2.2 days and <1 day for 3.94, 6.07 and 8.63 log10 FFU/mL CHIKV doses in the blood meal, respectively. DIR was inferred from experimental data for a larger set of CHIKV dose ranging from 3 to 8 log10 FFU/mL (Figure 3B). All the CHIKV doses tested led to 100% dissemination within the 40 days range used for predictions, although a longer time is required to reach this plateau at the lowest dose.

**Figure 3.**
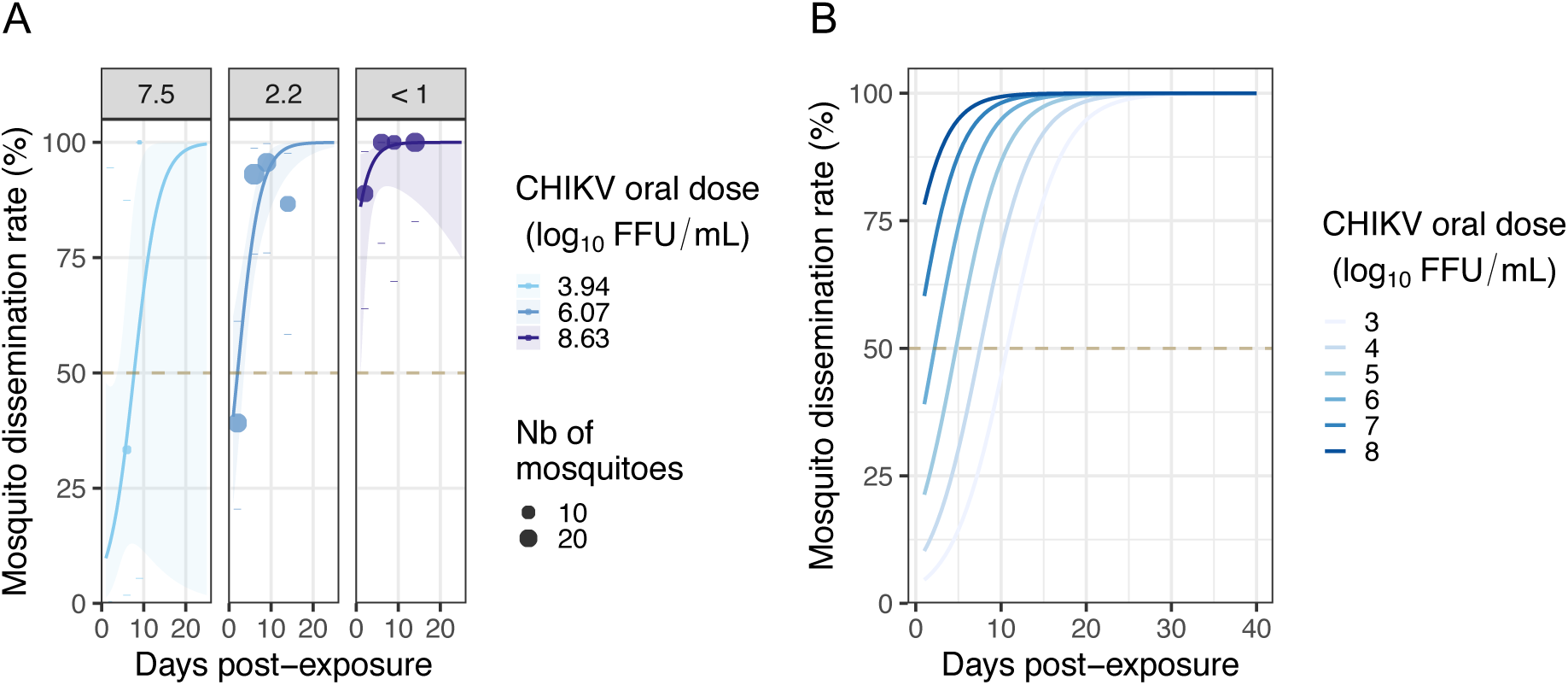
Dose-dependent dissemination rate of *Ae. albopictus* mosquitoes exposed to CHIKV 06.21. (A) The mosquito dissemination rate corresponds to the number of CHIKV-positive heads out of the infected (CHIKV-positive bodies) individuals, measured at 2, 6, 9 and 14 days post-exposure for three virus doses (3.94, 6.07 and 8.63 log10 FFU/mL) in the blood meal. Dot size is proportional to the number of mosquitoes tested. No disseminated females were detected at day 14 post-exposure at the 3.94 log10 FFU/mL dose. Logistic regression was used to model the time-dependent effect of the virus dose on mosquito dissemination rate. Lines correspond to fit values with their 95% confidence intervals displayed as ribbons. The time needed to reach 50 % dissemination is 7.5, 2.2 and <1 day for the 3.94, 6.07 and 8.63 log10 FFU/mL CHIKV doses, respectively, as indicated within each facet label. (B) Predicted dissemination dynamics according to virus dose and time post-exposure for a range of CHIKV blood meal titers (3 to 8 log10 FFU/mL). Refers to raw data table “Data_titer_EIPdyna_final.txt” (see data availability section).

### Time post-exposure modulates transmission rate and viral load in the saliva

The presence of infectious CHIKV particles in individual mosquito saliva collected by forced salivation technique was monitored at a fine time scale. This allowed us to measure the transmission rate (TR) and quantify individual viral load in saliva over time, and to estimate the extrinsic incubation period (EIP). Two virus doses (5.68 and 8.06 log10 FFU/mL) were used to obtain a workable proportion of infectious mosquitoes at a high number of time points that covers mosquito expected lifespan while remaining in the range of human viraemia. From day 2 post-exposure, TR increases following a sigmoid shape, reaching a plateau of around 60% for both doses (Figure 4A). TR was analysed using virus dose, time post-exposure and their interaction as explanatory variables, and only time was significant (Wald χ^2^, *P*time = 0.0037, *P*dose = 0.18, and *P*dose*time = 0.8). Infectious saliva samples were detected as soon as day 2 post-exposure to CHIKV, with a 33 % TR at dose 5.68 log10 FFU/mL (n=1/3 individuals tested) and 14 % at dose 8.06 log10 FFU/mL (n=2/14). The time needed to reach 50 % infectious mosquitoes (*i.e.* Extrinsic Incubation Period 50%, EIP50%) was 7.5 and 3.5 days for doses 5.68 and 8.06 log10 FFU/mL, respectively. A TR saturation at 100 % is a prerequisite to applying logistic regression analysis to the data. Therefore, the proportion of mosquitoes that would not ultimately transmit the virus was artificially removed from the dataset based on the predicted saturation level at each dose (*i.e*., 40 % of mosquitoes without the virus in their saliva were removed at each time post-infection). Logistic regression was used on these transformed data to predict TR across a range of doses over time (Figure S2).

**Figure 4.**
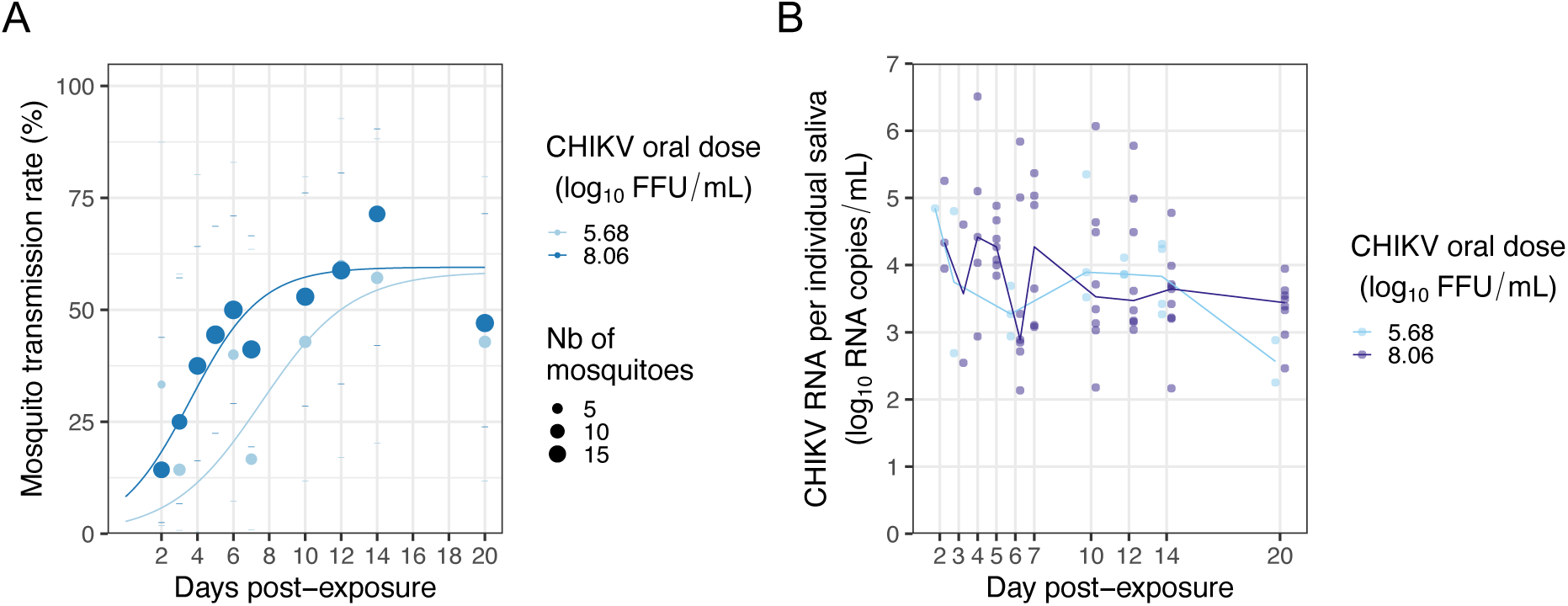
Transmission dynamics of CHIKV 06.21 by *Ae. albopictus*. (A) Mosquito transmission rate corresponds to the number of CHIKV-positive saliva out of the number of CHIKV-positive heads collected at 2, 3, 4, 5, 6, 7, 10, 12, 14, and 20 days post-exposure for two virus doses (5.68 and 8.06 log10 FFU/mL) in the blood meal. Dot size is proportional to the number of saliva tested. (B) CHIKV RNA load of each saliva scored positive for infectious CHIKV was measured by TaqMan RT-qPCR assay using a synthetic RNA as standard, then expressed in log10 CHIKV RNA copies/saliva. Each dot represents a saliva sample from a mosquito exposed to the indicated dose. Refers to raw data tables “Data_titer_EIPdyna_final.txt” and “Data_CHIKV_RNA_load_saliva.txt” (see data availability section).

To decipher if CHIKV load in the saliva could be associated with the virus dose mosquitoes were challenged with, total RNA was isolated at each time point from the saliva of individual mosquitoes exposed to 5.68 or 8.06 log10 FFU/mL. Viral load was measured by TaqMan RT-qPCR assay and analysed according to virus dose and time post-exposure. A high individual variation is noticed (up to 10,000 fold difference between individuals), although CHIKV load in the saliva seems to decrease over time (Figure 4B). Only the time post-exposure significantly affected viral load when considering saliva samples that were CHIKV-positive both in qRT-PCR and in infectious titration (Anova, *P*time = 0.006, *P*dose = 0.66 and *P*dose*time = 0.94). Of note, both time and virus dose affected viral load in the saliva when considering RNA-positive samples regardless of the presence of infectious virus (Figure S3).

### Simulation of CHIKV epidemic upon dose-dependent intra-vector dynamics

A stochastic agent-based model was used to assess the epidemiological impact of within-host CHIKV dynamics using the R package *nosoi*, as done previously for ZIKV (Lequime et al., 2020). Starting with one infected human in a population of susceptible humans and mosquitoes, the model simulates CHIKV transmissions according to human viraemia, its derived probability of mosquito infection, and virus transmission timeliness (EIP). The model was run 100 independent times for a maximum of 365 days for a range of eleven mean individual mosquito biting rates (1, 2, 3, 4, 5, 6, 7, 8, 9, 10 and 60 independent mosquitoes biting per person per day). Simulations led to large outbreaks (>100 secondary infections) even under a low mosquito biting rate (Figure 5A). The maximum threshold of mosquito infection was reached regardless of biting intensity, although the time needed to reach this threshold and the inter-simulation variation were higher at the lowest biting intensity (*i.e.* 1 bite per day) compare to other conditions (Figure 5B). Accordingly, secondary cases values distributions across simulations were narrow for all conditions except at 1 bite per day reflecting the explosive nature of the outbreak. The mean secondary case values increases as a function of the mosquito biting intensity with a mean (± SD) of 3.68 (± 1.92), 7.37 (± 2.71), 11.06 (± 3.32), 14.75 (± 3.84), 18.43 (± 4.29), 22.12 (± 4.7), 25.81 (± 5.07), 29.5 (± 5.43), 33.18 (± 5.75), 36.87 (± 6.07) and 221.15 (± 6.09) for 1, 2, 3, 4, 5, 6, 7, 8, 9, 10 and 60 mosquito bites per person per day, respectively (Figure 5C).

**Figure 5.**
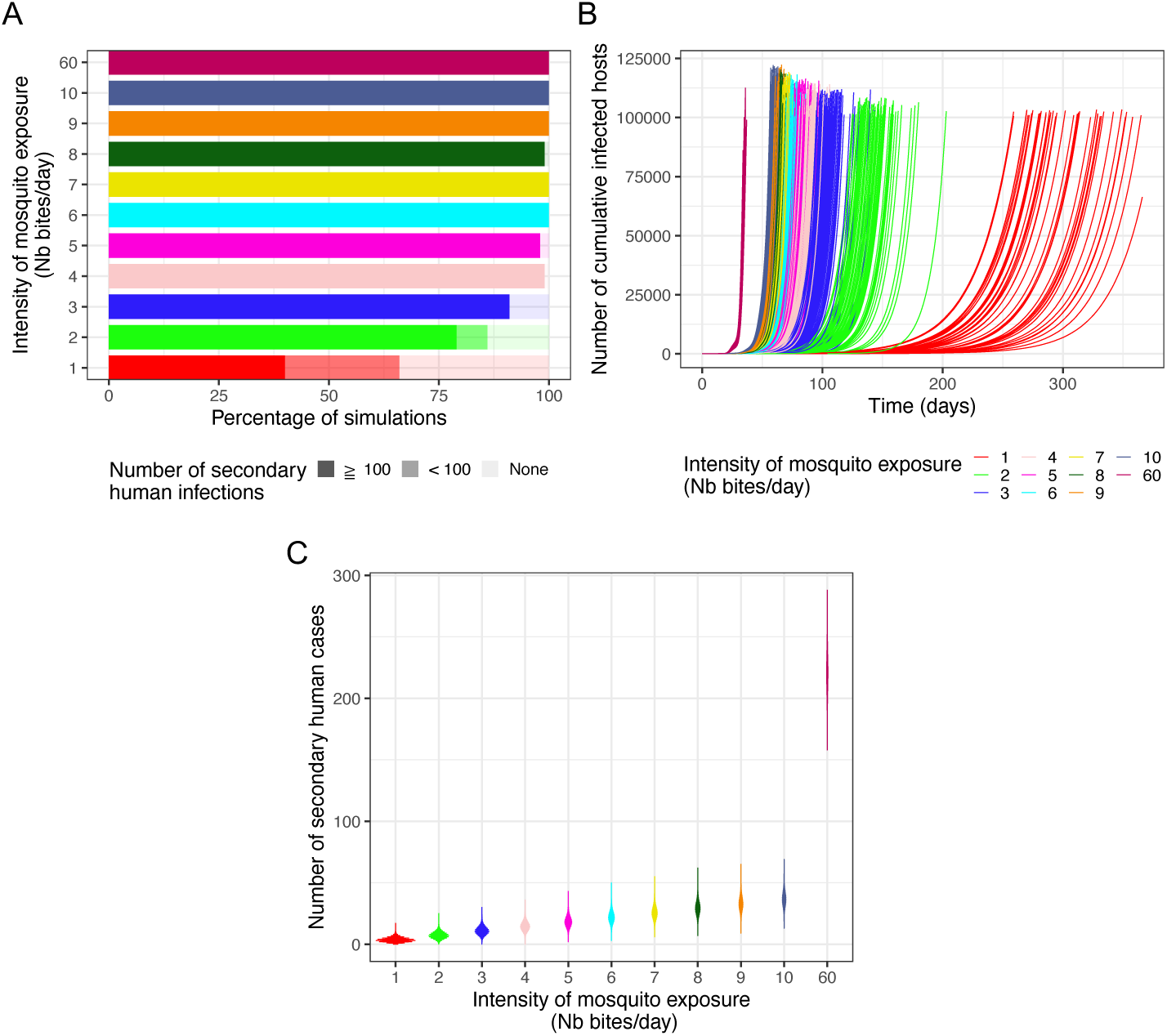
Influence of dose-dependent intra-mosquito CHIKV dynamics on outbreak simulations with various levels of mosquito bites. Stochastic agent-based epidemiological simulations considering within-vector infection dynamics on transmission probability during mosquito-human infectious contacts were performed in 100 independent replicates. A total of 11 mosquito bite intensity levels were tested: 1, 2, 3, 4, 5, 6, 7, 8, 9, 10 and 60 bites per human per day. (A) Stacked proportions of outbreak simulations resulting in no secondary infected human host, < 100 and ≧ 100 infected human hosts. (B) Cumulative number of infected humans over time. Each curve represents a simulation run. (C) Violin plots showing the number of secondary cases densities for each intensity of mosquito exposure. Refers to raw data tables “Compiled_results_run5.csv”, “cumulative_run5.csv” and “R0dist_run5.csv” (see data availability section).

## Discussion

Vector competence (VComp) of *Aedes albopictus* for chikungunya virus (CHIKV) has been widely studied, notably since the La Réunion outbreak in 2006. Studies outlined a strong impact of mosquito and CHIKV genotype as well as temperature on *Ae. albopictus* potential for CHIKV transmission, including large VComp variations among worldwide *Ae. albopictus* populations exposed to the highly transmissible La Réunion 2006 CHIKV 06.21 isolate (Zouache et al., 2014; Mariconti et al., 2019; Gloria-Soria et al., 2020; Vega-Rúa et al., 2020). However, knowledge gaps remain regarding intra-vector virus dynamics and its impact on CHIKV epidemic potential notably regarding the virus dose. Our work contributes to fill these gaps, providing important data on the interplay between CHIKV and *Ae. albopictus* as well as on the vireamia-dependent human infectiousness for mosquitoes.

### Dose and time-dependent barriers to CHIKV intra-vector dynamics

*Ae. albopictus* midgut infection is strongly influenced by CHIKV dose in the blood meal. Previous studies documented an oral infectious dose for 50 % of the mosquitoes (OID50%) ranging from 1.7 to 3.52 log10 infectious particles per mL of blood for CHIKV 06.21 and thus lower than the 5.60 log10 FFU/mL OID50% estimated in our *Ae. albopictus* population (Tsetsarkin et al., 2007; Pesko et al., 2009; Hurk et al., 2010). ZIKV OID50% was 5.62 log10 FFU/mL in *Ae. albopictus* while in *Ae. aegypti*, ZIKV and DENV OID50% ranged from 4.73 to 8.10 log10 FFU/mL and from 3.95 to 5.5 log10 PFU/mL, respectively (Nguyet et al., 2013; Aubry et al., 2020; Lequime et al., 2020). The CHIKV time-independent infection rate and relatively low OID50% in *Ae. albopictus* suggests that primary infection of midgut cells is rapid and efficient. Such CHIKV midgut infection pattern could be promoted by the presence of several potential midgut receptors for *Alphavirus* entry in mosquitoes (Franz et al., 2015). Importantly, a 0.5 log10 FFU/mL increase in OID50% during the exponential phase results in twice as much infected mosquitoes, which could exacerbate outbreaks, notably upon large vector densities. Therefore, dose-response experiments at a small dose range for each virus and mosquito genotype of interest would significantly improve our understanding of vector competence.

According to our results, CHIKV dissemination from the *Ae. albopictus* midgut depends on the interaction between time post-exposure and virus dose. A previous study showed that at day 6 post-exposure, CHIKV dissemination rate in *Ae. albopictus* increases with virus dose, being ∼10%, ∼50% and >80% upon 3.6, 4.4 and 5.2 log10 PFU/mL in the blood meal respectively (Pesko et al., 2009). Here, we show that if at least 6 days are needed to reach ∼30 % dissemination upon 3.94 log10 FFU/mL, virus doses of 6.07 and 8.63 log10 FFU/mL led overall to ∼90 % dissemination regardless of the time point (except at day 2 for the 6.07 log10 FFU/mL dose where only 40 % dissemination was observed). Of note, CHIKV dissemination rate at 3.94 log10 FFU/mL shall be interpreted with caution due to the low sample size that arise directly from the low infection rate (1.9 to 6.5%, n = 37 to 53 individuals per time point). These results reveal the ability of CHIKV to efficiently disseminate from the midgut, as modelling for a larger dose range estimates that all infected mosquitoes eventually disseminate even at the lowest dose considered (2 log10 FFU/mL). CHIKV and ZIKV present a nearly identical OID50% suggesting similar midgut infection potential in *Ae. albopictus*. However, ZIKV dissemination is slower and, to a minor extend, reaches lower value compared to CHIKV (Lequime et al., 2020). This discrepancy might be due to viral replication in the midgut as dissemination rate correlates with midgut viral load (Houk et al., 1981; Bosio et al., 1998; Dickson et al., 2014; Vazeille et al., 2019; Carpenter et al., 2021). CHIKV dissemination might arise from an efficient replication in the midgut tissue. Recently, the CHIKV 3’ untranslated region was recently shown to promote dissemination through an increased viral replication in the mosquito midgut (Merwaiss et al., 2020).

Ultimately, arboviruses infect and replicate in mosquito salivary glands, this step being essential to allow virus transmission to the host (Vega-Rúa et al., 2015; Raquin & Lambrechts, 2017). Virus prevalence in the head is often used as a proxy for transmission potential but it is likely an overestimate due to salivary glands barriers, notably for CHIKV (Sanchez-Vargas et al., 2021). Our study shows that ∼60 % of the mosquitoes with disseminated infection eventually become infectious, this being an underestimate of mosquito-to-host transmission potential due to the use of forced salivation technique (Gloria-Soria et al., 2022). Moreover, transmission rate strongly depends on the time post-exposure. Previously, the time to reach 50 % of infectious mosquitoes in the population (EIP50%) was estimated by a meta-analysis at 7 days (± 1 day), based on dissemination data and for mosquitoes exposed to relatively high virus doses (Christofferson et al., 2014). Despite no overall effect of virus dose on the transmission rate (*P*dose = 0.18), CHIKV EIP50% was 7.5 and 3.5 days in mosquitoes exposed to 5.68 or 8.06 log10 FFU/mL, respectively, suggesting that virus dose might influence CHIKV transmission. Increasing sample size and/or testing an intermediate virus dose (*e.g.* 6 log10 FFU/mL) could help to better capture the impact of virus dose on transmission rate and resolve this discrepancy. Interestingly, when considering all CHIKV-positive salivas (including the ones with only CHIKV RNA but found negative during infectious titration), the CHIKV RNA load in the saliva depends on time post-exposure and virus dose but not in interaction. Overall, the CHIKV load in saliva seems higher at high dose but decreases overtime, questioning arbovirus-salivary glands interaction. In a previous study, individual ZIKV-disseminated *Ae. aegypti* mosquitoes were offered successive non-infectious blood meals in an attempt to monitor expelled virus during feeding in a non-sacrificial manner. Authors observed an on/off presence of ZIKV in the blood meal for a single individual over time; however, whether this is due to biological or methodological causes is unclear (Mayton et al., 2021). In *Ae. albopictus*, up to 10,000-fold difference of CHIKV RNA load in the saliva was found between individuals at a given time point and dose, in accordance with previous results (Dubrulle et al., 2009; Bohers et al., 2020; Robison et al., 2020). No correlation was found between CHIKV titer in the salivary glands and in the saliva, that might be linked with such inter-individual variations (Sanchez-Vargas et al., 2019). Despite several studies that identified histological and genetical factors modulating viral infection in this tissue, it is still unclear how and in which amount infectious virions are produced in salivary glands and then transferred into the saliva over time (Ciano et al., 2014; Modahl et al., 2019; Chowdhury et al., 2021; Sanchez-Vargas et al., 2021). Notably, viral particles in the saliva might use specific viral factors and/or mosquito saliva proteins to persist in the saliva and promote their transmission (Pompon et al., 2017; Marin-Lopez et al., 2021). This is key as virus titer in the mosquito inoculum is associated with viraemia level and symptoms severity in mice and macaques models (Labadie et al., 2010; Zhang et al., 2022).

### Genetic and environmental factors impacting intra-vector dynamics

VComp is a composite phenotype that also depends on the interaction between virus genotype, mosquito genotype and temperature (Zouache et al., 2014). From a virus perspective, some CHIKV mutations that impact VComp were already described and could be useful for epidemiological monitoring, even if data suggest that overall CHIKV swarm maintains an intermediate mutation frequency to avoid fitness loss in the mosquito (Coffey et al., 2014). Transcriptomics and quantitative genetics studies led to the identification of mosquito genetic loci that constitute interesting targets towards engineered vector control approaches, as shown for *Flaviviridae* (Bosio et al., 2000; Raquin et al., 2017; Aubry et al., 2020; Merkling et al., 2020; Williams et al., 2020; Dong et al., 2022). In addition, mosquitoes host a microbiota composed of bacteria, viruses, fungi and protists that have a major impact on vector’s biology (Guégan et al., 2018). Some of these micro-organisms are associated with a decrease in arbovirus transmission and constitute interesting vector control tools, like *Wolbachia* or *Delftia tsuruhatensis* bacteria blocking DENV and *Plasmodium* infection, respectively or insect-specific viruses modulating *Ae. aegypti* vector competence for DENV (Olmo et al., 2018; 2023; Huang et al., 2023). Despite an antiviral activity of *Wolbachia* against CHIKV in *Ae. aegypti*, as well a in *Ae. albopictus* C6/36 cell line, no CHIKV blocking was detected in *Ae. albopictus* mosquitoes (Mousson et al., 2012; Raquin et al., 2015; Aliota et al., 2016). Moreover, *Ae. albopictus* infection by CHIKV impacts mosquito bacterial community composition while several *Ae. albopictus* symbionts were associated with an increase of CHIKV infection (Zouache et al., 2012; Monteiro et al., 2019). The reason for this lack of microbiota-mediated antiviral blocking against CHIKV in *Ae. albopictus* remains obscure, but could depend on mosquito and virus genotype and/or temperature as *Ae. albopictus* microbiota composition depends on the temperature (Bellone et al., 2023). This interaction should be further studied as it could impact arbovirus transmission, as suggested by models estimating that the release of *Wolbachia*-infected *Ae. aegypti* against DENV will be less efficient upon long heatwaves due to loss of *Wolbachia* infection upon high temperatures (Vásquez et al., 2023). With the exception of DENV genotype (Fontaine et al., 2018), the impact of aforementioned factors was not tested on VComp dynamics and it will be interesting to determine if these factors, beyond modulating VComp at discrete time, impact the proportion of infectious mosquitoes over time.

### Human viremia and infectiousness to mosquitoes

A large majority of VComp studies exposed *Ae. albopictus* to a high (>7 log10 plaque-forming units (PFU)/mL) CHIKV titer in the blood meal (Coffey et al., 2014). According to our estimate, CHIKV viraemia in humans can reach >7 log10 PFU equivalent per mL of blood, although this corresponds to the highest value measured within a short time window (<2 days). Our CHIKV viraemia estimate from human longitudinal data lasts from 4 to 12 days with a mean maximum titer of 7.55 log10 PFU/mL, which is largely supported by previous human and nonhuman studies (Lanciotti et al., 2007; Panning et al., 2008; Labadie et al., 2010; Schwartz & Albert, 2010). The mean human viraemia is above 3.94 log10 FFU/mL during 5.5 days, implying that humans are infectious to mosquitoes during more than half of their viraemia. These data are key to improve sanitary guidelines for the management of CHIKV infections and spread. However, major differences in CHIKV viraemia magnitude and length are observed between individual hosts, which can be associated with host and/or virus genotypes as observed in dengue virus (DENV)-infected patients (Nguyet et al., 2013). In addition, if artificial mosquito blood feeding limits variation in blood composition and promotes reproducibility, it does not necessary represent the native infectiousness of human-derived arbovirus for mosquitoes. This is partly linked to host plasma factors level (IgM, IgG, low-density lipoproteins or gamma-aminobutyric acid) while asymptomatic DENV cases are more infectious to mosquitoes than symptomatic counterparts at a given dose suggesting a link between host immune response and vector transmission (Nguyet et al., 2013; Duong et al., 2015; Wagar et al., 2017; Zhu et al., 2017). Moreover, for CHIKV, ∼85 % infections are symptomatic with a median blood titer about 100-fold higher compare to asymptomatic carriers, although this difference was not statistically significant (Appassakij et al., 2013). Thus, improving epidemiological models by implementing the time-dependant human host infectiousness to mosquitoes represents an interesting lead to better anticipate and prevent CHIKV outbreaks. This notably prompts the need for viraemia monitoring over time in large groups of patients infected by arboviruses, using standardized virus titration procedures to facilitate comparisons and calibrate dose-response experiments in mosquitoes. This also requires other improvements of modelling strategies to account for mosquito and human populations structure, sanitary measures as well as *Ae. albopictus* tendency to take several consecutive blood meals. Indeed, virus dissemination increases with gonotrophic cycles and that successive bloodmeals are associated with a shortened EIP (Delatte et al., 2010; Armstrong et al., 2019; Fikrig & Harrington, 2021; Mulatier et al., 2023). Although those limitations could modulate the explosiveness of the outbreak and could be improved, our data support that CHIKV transmission potential of local *Ae. albopictus* is not a limiting factor for local CHIKV emergence and spread. Finally, such a standardized experimental design can be used to investigate the impact of additional (a)biotic factors on VComp dynamics.

Beyond virus dose and time post virus exposure, mosquito and virus genotype, mosquito microbiota and temperature are currently identified as major VComp drivers. Therefore, locally-acquired VComp data from area at risk for arbovirus circulation are needed. This could be achieved by exposing autochthonous field-derived mosquito populations to virus strains currently circulating (or at risk of introduction) upon a range of virus dose and temperature spanning the human viremia and the mosquito season, respectively, while controlling experimentally mosquito microbiota. This is unlikely to be done within a single experiment but will require the incremental acquisition of data sets for each factor, underlying the importance of experimental procedure standardization. Interestingly, modelisation of VComp based on available data could help to target a range of the factor’s values to be tested experimentally, as exemplified with temperature (Shocket et al., 2020). These factors act together in interaction making difficult to dissociate their real impact on VComp in the field. Deciphering the complex interplay between each factor on VComp is challenging but feasible (Audsley et al., 2017) and holistic interaction studies could help to address this issue experimentally (Brinker et al., 2019). Beyond experimental conditions, our work underlines that monitoring intra-vector dynamics rather than end-point VComp is key to accurately quantify VComp variations and better estimate VCap (Christofferson & Mores, 2011). Several studies estimated VCap through the lens of mosquito density upon environmental variations (*e.g.* temperature, micro-climate, land cover) but with limited ecological (mosquito survival rate, biting rate, density per host) and VComp data, notably regarding intra-vector dynamics (Murdock et al., 2017; Wimberly et al., 2020; Peña-García et al., 2023). This highlights the current need for additional VComp and VCap-related studies using field-derived material, as well as increasing efforts between vector biology and modeling fields towards an integrative VCap estimation notably regarding intra-vector arbovirus dynamics. Altogether, this will improve vector control strategies and case management by health authorities.

## Appendices

FigS1 - CHIKV infectious titer is stable upon a one hour incubation at 37°C in human erythrocytes suspension. Refers to raw data table “blood_titration_FFU.txt” (see data availability section).

FigS2 - Rescaled mosquito transmission dynamics. Refers to raw data table “Data_titer_EIPdyna_final.txt” (see data availability section).

FigS3 - Time-course of CHIKV load in mosquito saliva. Refers to raw data table “Data_CHIKV_RNA_load_saliva.txt” (see data availability section).

TabS1 - Proportion of infected, disseminated and infectious mosquitoes over time according to the dose of CHIKV in the blood meal. Refers to raw data table “Raw_data_viginier_et_al2023” (see data availability section).

## Supporting information

Viginier_et_al_supp_https://doi.org/10.24072/pci.infections.100091

## Data Availability

All data produced are available online at https://doi.org/10.5281/zenodo.8033668

## Acknowledgements

This project was funded by the scientific breakthrough project Micro-Be-Have (Microbial impact on insect behavior) of the Université de Lyon within the program Investissements d’Avenir (ANR-11-IDEX-0007; ANR-16-IDEX-0005). We thank the Equipex InfectioTron program and its project manager Isabelle Weiss. We also thank all the members of the Micro-Be-Have consortium for insightful discussions. We thank Anna-Bella Failloux and Patrick Mavingui for the CHIKV 06.21 isolate. We acknowledge the contribution of SFR Biosciences (UAR3444/CNRS, US8/Inserm, ENS de Lyon, Universite Claude Bernard Lyon 1) AniRa biosafety level 3 platform (Marie-Pierre Confort) and plateau Analyse Génétique et Cellulaire (Bariza Blanquier) facilities in Lyon. We also thank 2 anonymous reviewers for helpful comments on a previous version of this manuscript.

## Data, scripts, code, and supplementary information availability

Data, R scripts, supplementary information and main figures in full size are available online: https://doi.org/10.5281/zenodo.8033668

## Conflict of interest disclosure

The authors declare that they comply with the PCI rule of having no financial conflicts of interest in relation to the content of the article.

Sebastian Lequime is a recommender for PCI infections.

## Author contributions

**BV**: Investigation; Data curation; Validation **LC**: Investigation; Data curation; Validation **CG**: Investigation; Data curation; Validation **EM**: Resources

**CM**: Resources

**CVM**: Funding acquisition; Resources; Supervision; Writing – review and editing

**GM**: Funding acquisition; Writing – review and editing

**AF**: Data curation; Formal analysis; Software; Visualization; Methodology; Writing – review and editing

**SL**: Data curation; Formal analysis; Software; Visualization; Methodology; Writing – review and editing

**MR**: Funding acquisition; Supervision; Writing – review and editing

**FA**: Funding acquisition; Project administration; Supervision; Writing – review and editing

**VR**: Conceptualization; Investigation; Formal analysis; Data curation; Methodology; Investigation; Project administration; Supervision; Validation; Visualization; Writing – original draft; Writing – review and editing.

