## Supplementary material for "Chikungunya intra-vector dynamics in *Aedes albopictus* from Lyon (France) upon exposure to a human viremia-like dose range reveals vector barrier’s permissiveness and supports local epidemic potential": Viginier_et_al_supp_https://doi.org/10.24072/pci.infections.100091

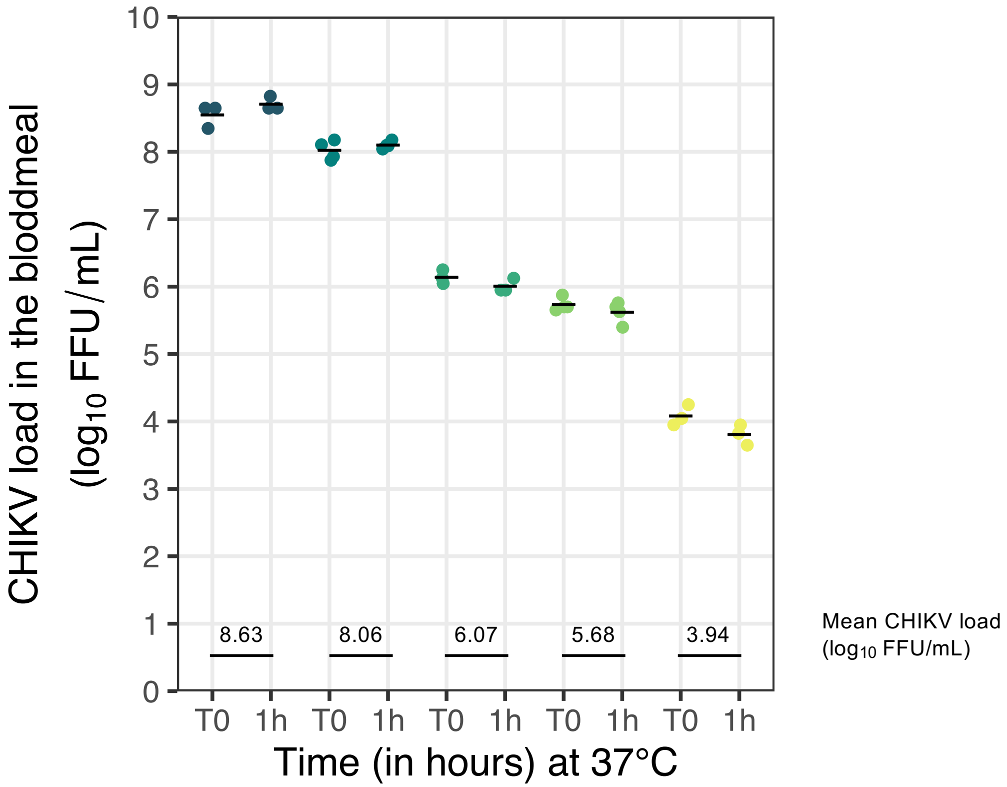


Figure S1 - CHIKV infectious titer is stable upon a one hour incubation at 37°C in human erythrocytes suspension. Blood meal aliquots were harvested immediately after mixing virus suspension with washed human erythrocytes (T0) and after one hour at 37°C in Hemotek artificial feeding system (1h). Each targeted dose is indicated by a different color. One or two aliquots per dose and time (T0 and 1h) were titrated by fluorescent focus assay (in technical triplicate or duplicate, respectively) to determine CHIKV infectious titer in FFU/mL. Mean infectious titers between T0 and 1h for each dose are indicated.

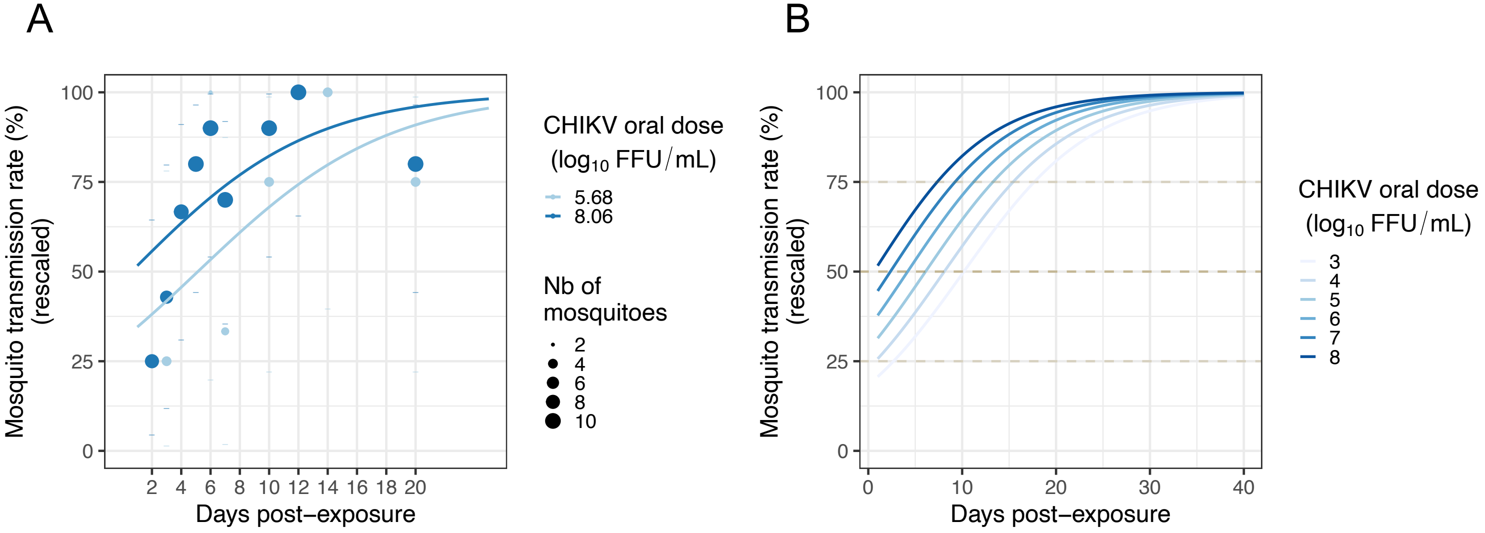


Figure S2 - (A) Rescaled mosquito transmission dynamics. Each dot corresponds to the proportion (in %) of *Ae. albopictus* female saliva positive for CHIKV infection (as determined by detection of infectious virus) at ten time points (2, 3, 4, 5 , 6, 7, 10, 12, 14, and 20 days post-exposure) for two virus doses (5.68 and 8.06 $log_{10}$ FFU/mL) in the blood meal. The dot size is proportional to the number of saliva tested. (**B**) Based on panel B, predicted transmission dynamics according to virus dose and time post-exposure for a range of CHIKV blood meal titers (3 to 8 $log_{10}$ FFU/mL).

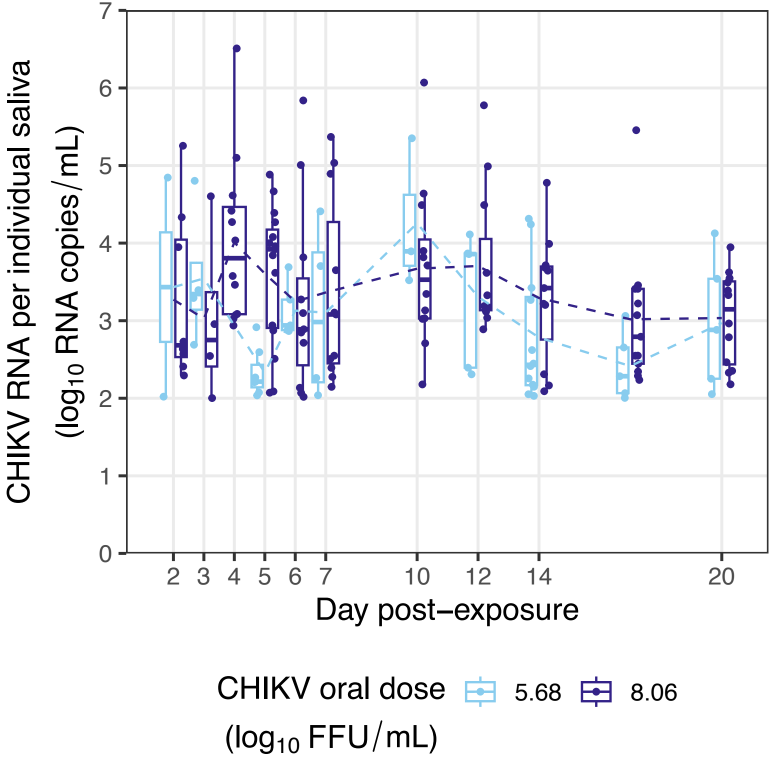


Figure S3 - Time course of CHIKV load in mosquito saliva. CHIKV RNA load in each saliva, including samples without infectious CHIKV, was measured by TaqMan RT-qPCR assay using a synthetic RNA as standard and then expressed in $log_{10}$ RNA copies/mL. Each dot represents a saliva sample from a mosquito exposed to the indicated virus dose (in $log_{10}$ FFU/mL).

| experiment | dose(log10FFUpermL) | daype | IR | IR_CI_low | IR_CI_up | IR_sampleSize | DR | DR_CI_low | DR_CI_up | DR_sampleSize | TR | TR_CI_low | TR_CI_up | TR_sampleSize |
|---|---|---|---|---|---|---|---|---|---|---|---|---|---|---|
| exp1 | 3.94 | 2 | 1.886792 | 0.09856963 | 11.37973 | 53 | 0 | 0 | 94.53792 | 1 | NA | NA | NA | NA |
| exp1 | 3.94 | 6 | 6.521739 | 1.69956551 | 18.92926 | 46 | 33.33333 | 1.765279 | 87.46655 | 3 | NA | NA | NA | NA |
| exp1 | 3.94 | 9 | 2.702703 | 0.14124326 | 15.80945 | 37 | 100 | 5.462076 | 100 | 1 | NA | NA | NA | NA |
| exp1 | 3.94 | 14 | 0 | 0 | 12.31534 | 35 | NA | NA | NA | 0 | NA | NA | NA | NA |
| exp1 | 6.07 | 2 | 67.647059 | 49.37124696 | 82.02451 | 34 | 39.13043 | 20.467675 | 61.21749 | 23 | NA | NA | NA | NA |
| exp1 | 6.07 | 6 | 85.294118 | 68.16538146 | 94.45638 | 34 | 93.10345 | 75.788826 | 98.7964 | 29 | NA | NA | NA | NA |
| exp1 | 6.07 | 9 | 65.714286 | 47.73795578 | 80.31557 | 35 | 95.65217 | 76.032219 | 99.77262 | 23 | NA | NA | NA | NA |
| exp1 | 6.07 | 14 | 75 | 50.58845414 | 90.40674 | 20 | 86.66667 | 58.38899 | 97.6562 | 15 | NA | NA | NA | NA |
| exp1 | 8.63 | 2 | 94.736842 | 71.89258903 | 99.72465 | 19 | 88.88889 | 63.926817 | 98.05172 | 18 | NA | NA | NA | NA |
| exp1 | 8.63 | 6 | 100 | 78.12431493 | 100 | 18 | 100 | 78.124315 | 100 | 18 | NA | NA | NA | NA |
| exp1 | 8.63 | 9 | 100 | 69.87459609 | 100 | 12 | 100 | 69.874596 | 100 | 12 | NA | NA | NA | NA |
| exp1 | 8.63 | 14 | 100 | 82.82849932 | 100 | 24 | 100 | 82.828499 | 100 | 24 | NA | NA | NA | NA |
| exp2 | 5.5 | 2 | NA | NA | NA | NA | NA | NA | NA | NA | 33.33333 | 1.7652794 | 87.46655 | 3 |
| exp2 | 5.5 | 3 | NA | NA | NA | NA | NA | NA | NA | NA | 14.28571 | 0.7502816 | 57.99217 | 7 |
| exp2 | 5.5 | 4 | NA | NA | NA | NA | NA | NA | NA | NA | 0 | 0 | 80.21325 | 2 |
| exp2 | 5.5 | 6 | NA | NA | NA | NA | NA | NA | NA | NA | 40 | 7.258404 | 82.95764 | 5 |
| exp2 | 5.5 | 7 | NA | NA | NA | NA | NA | NA | NA | NA | 16.66667 | 0.8762291 | 63.51774 | 6 |
| exp2 | 5.5 | 10 | NA | NA | NA | NA | NA | NA | NA | NA | 42.85714 | 11.8083011 | 79.76283 | 7 |
| exp2 | 5.5 | 12 | NA | NA | NA | NA | NA | NA | NA | NA | 60 | 17.0423581 | 92.7416 | 5 |
| exp2 | 5.5 | 14 | NA | NA | NA | NA | NA | NA | NA | NA | 57.14286 | 20.2371694 | 88.1917 | 7 |
| exp2 | 5.5 | 20 | NA | NA | NA | NA | NA | NA | NA | NA | 42.85714 | 11.8083011 | 79.76283 | 7 |
| exp2 | 8.0 | 2 | NA | NA | NA | NA | NA | NA | NA | NA | 14.28571 | 2.5139354 | 43.84934 | 14 |
| exp2 | 8.0 | 3 | NA | NA | NA | NA | NA | NA | NA | NA | 25 | 6.6935697 | 57.16439 | 12 |
| exp2 | 8.0 | 4 | NA | NA | NA | NA | NA | NA | NA | NA | 37.5 | 16.2837034 | 64.12641 | 16 |
| exp2 | 8.0 | 5 | NA | NA | NA | NA | NA | NA | NA | NA | 44.44444 | 22.4047514 | 68.65307 | 18 |
| exp2 | 8.0 | 6 | NA | NA | NA | NA | NA | NA | NA | NA | 50 | 29.0310215 | 70.96898 | 18 |
| exp2 | 8.0 | 7 | NA | NA | NA | NA | NA | NA | NA | NA | 41.17647 | 19.4278913 | 66.5465 | 17 |
| exp2 | 8.0 | 10 | NA | NA | NA | NA | NA | NA | NA | NA | 52.94118 | 28.533856 | 76.14276 | 17 |
| exp2 | 8.0 | 12 | NA | NA | NA | NA | NA | NA | NA | NA | 58.82353 | 33.4535019 | 80.57211 | 17 |
| exp2 | 8.0 | 14 | NA | NA | NA | NA | NA | NA | NA | NA | 71.42857 | 42.0031729 | 90.41824 | 14 |
| exp2 | 8.0 | 20 | NA | NA | NA | NA | NA | NA | NA | NA | 47.05882 | 23.8572447 | 71.46614 | 17 |

Table S1 - Proportion of infected, disseminated and infectious mosquitoes over time according to the dose of CHIKV in the blood meal. For each experiment (exp1 and exp2), the virus dose (in $\log_{10}$ FFU/mL of blood) and the day post-exposure (daype) are indicated. The infection rate (IR) corresponds to the number of CHIKV-positive bodies out of the number of blood-fed females, with the lower (IR_CI_low) and upper (IR_CI_up) limits of the 95% confidence interval and the number of tested mosquitoes (IR_sampleSize). The same indicators are provided for the dissemination rate (DR) and transmission rate (TR) that correspond to the number of CHIKV-positive heads out of the number of CHIKV-positive bodies and the number of CHIV-positive saliva out of the number of CHIKV-positive heads, respectively. NA's indicate conditions for which samples were not harvested or that could not be tested.
